# Eating Disorders Spectrum during COVID Pandemic: a systematic review

**DOI:** 10.1101/2021.04.16.21255390

**Authors:** Mario Miniati, Francesca Marzetti, Laura Palagini, Donatella Marazziti, Graziella Orrù, Ciro Conversano, Angelo Gemignani

**Author notes:** **Correspondence:** Mario Miniati.

## Abstract

**Background:** Several data suggest that COVID-19 pandemic might exacerbate or trigger Eating Disorders (EDs). The aim of this paper was to summarize present literature on COVID pandemic and EDs.

**Methods:** Literature search, study selection, methods, and quality evaluation were performed following PRISMA Guidelines.

**Results:** The systematic search permitted the identification of 91 studies; 21 papers were eligible and included in the review. Nine papers (42.9%) evaluated the effect of pandemic and associated protective and risk factors in EDs patients, ten (47.6%) explored the prevalence of disturbed eating behaviours and risk factors for exacerbating EDs in the general population, and the remaining two (9.5%) were qualitative studies describing the impact of lockdown and quarantine on EDs patients.

Their analysis revealed five main findings: 1) changes in physical activities routines were related to a worsening of preoccupation on weight/body shape; 2) food access limitation during pandemic represented a risk factors for both triggering and exacerbating EDs; 3) restriction in healthcare facilities contributed to increase anxiety levels and modifies treatment compliance; 4) social isolation was related to symptoms’ exacerbation in EDs patients who are home-confined with family members; 5) conflicts and difficulties in relationships with ‘no way out’ were maintenance factors for EDs symptoms, especially in adolescents and young adults.

**Conclusion:** COVID-19 pandemic had a negative impact on EDs that might be triggered or worsened by the exceptional conditions deriving from COVID-19-related stress in predisposed subjects. Patients already affected by EDs experienced a worsening of their clinical conditions and related quality of life.

## 1 INTRODUCTION

Changes in everyday activities and in lifestyle due to SARS-COV2 disease 2019 (COVID-19) pandemic and related quarantine measures heavily impacted mental health of the general population (Marazziti, Pozza, Di Giuseppe, & Conversano, 2020; Poli, Gemignani, & Conversano, 2020) and of special populations (Conversano, Marchi, & Miniati, 2020; Orrù et al., 2021). These changes may represent an additional burden for both individuals with a pre-existing eating disorders (EDs) and those from the general population predisposed to develop them belonging into the so-called ‘*anorexia-bulimia spectrum*’ (Miniati & Marazziti, 2019). A series of factors may have detrimental impacts on psychological wellbeing, eating habits and EDs onset or recovery, including: disruption to living situations, social distancing restrictions, difficult access to healthcare, and societal changes to food behaviours and technology usage (Ammar et al., 2020). Therefore, at times of uncertainty and instability, ED symptoms may increase as their functions can be providing control and/or safety. Psychosocial stressors stemming from COVID-19 pandemic and resulting stay-at-home confinement, may exacerbate ED-related triggers and represent a challenging environment for individuals with anorexia nervosa (AN), bulimia nervosa (BN) and binge-eating disorder (Hensley, 2020; McMenemy, 2020). Amongst patients with anorexia nervosa (AN), the hypertrophic sense of control and the intense polarization on body weight and shape might worsen, when self-control seems altered by the impact of exceptional external factors, such as those occurring during COVID pandemic and lockdown (Schlegl, Maier, Meule, & Voderholzer, 2020). Again, they may foster skipping meals and restricting calories or, conversely, they may increase binge eating due to the availability of food at home brought about by food insecurity and hoarding of food (Touyz, Lacey, & Hay, 2020; Weissman, Bauer, & Thomas, 2020). Moreover, preliminary observations focused on the risk to develop an ED in patients who contracted COVID-19, renewing questions on the role of immunologic and neurobiological factors as potential triggers for AN and BN through inflammatory processes, together with behavioural changes induced by the infection, such as loss of appetite and decrease in food intake (Breithaupt, Kholer-Forsberg, Larsen et al., 2019). A vulnerable patients’ population such as the one represented by EDs is at high risk of acute and long-term consequences from COVID-19 pandemic, while also considering the relevant restrictions in psychiatric and psychological services and the access limitations to both inpatients and outpatients facilities.

Aim of this paper is to summarize and comment on present literature on the impact of COVID pandemic on EDs spectrum.

## 2 MATERIALS AND METHODS

We adhered to the Preferred Reporting Items for Systematic Review and Meta-Analyses (PRISMA) guidelines in completion of this systematic review (Moher, Liberati, Tetzlaff, & Altman, 2009).

### 2.1 Protocol and registration

This systematic review is not included in a research protocol.

### 2.2 Eligibility Criteria

We included all studies published between 2019 and October 2020, by using PubMed, SCOPUS and Google Scholar, provided that they met the following criteria: 1) written in English; 2) original articles on studies with a longitudinal design; and 3) prospective or retrospective, observational (analytical or descriptive), experimental or quasi-experimental, controlled or non-controlled studies. Reviews and non-original articles (i.e., case reports, editorials, letters to the Editor and book chapters) were not included. We found no RCTs or long-term follow-up studies derived from RCTs, therefore, an assessment of risk of bias of individual studies was not performed.

### 2.3 Information Source and Strategy

The literature search was designed and performed independently in duplicate by two authors. The PubMed database was systematically screened using the following terms: [EATING DISORDER] AND [COVID] OR [LOCKDOWN] (n=48); [COVID] AND [BINGE-EATING] (n=16); [COVID] AND [BULIMIA NERVOSA] (n=16); [COVID] AND [ANOREXIA NERVOSA] (n=8), leading to a total of 88 papers. Additional three papers were identified after a second manual search was carried out to retrieve other papers that had not been identified with the initial strategy and included, leading to a total of 91 papers. After analyzing their titles and abstracts, according to the eligibility criteria, 21/91 (23.1%) papers were chosen and included in the final sample, whereas 70/91 papers (76.9%) were excluded for the following reasons: 34 papers (34/91; 37.4%) were removed as duplicated records; 31 papers (31/; 34.1%) were excluded because not pertinent to the selected topic; 5/91 (5.5%) papers were excluded because not in English (see **Fig. 1** for details).

**Figure 1.**
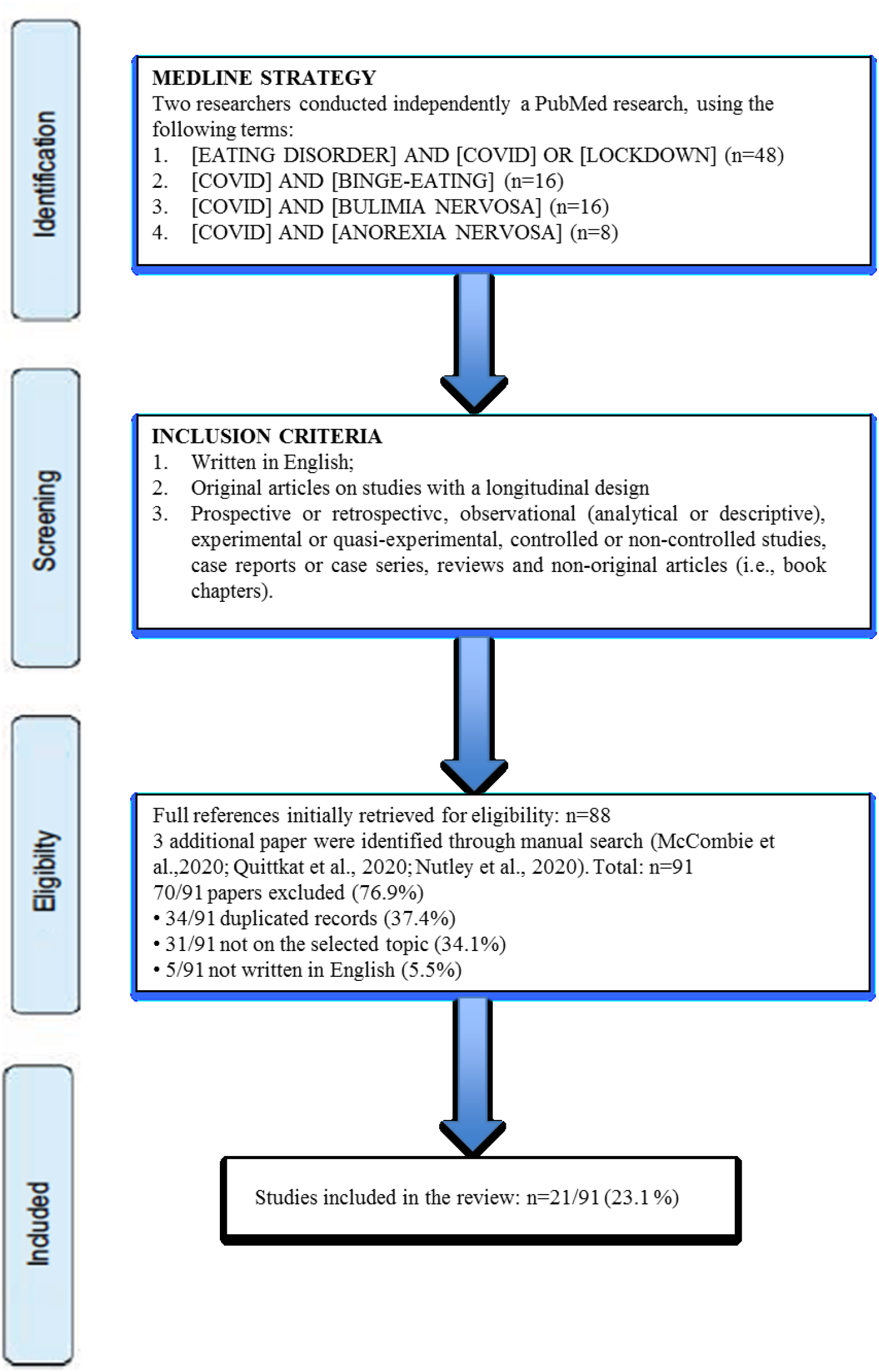
PRISMA flow diagram of studies’ selection.

### 2.4 Study selection

Two authors (MM and FM) independently screened the resulting articles for their methodology and appropriateness for inclusion. Consensus discussion was used to resolve reviewers’ disagreements.

### 2.5 Data collection process and data items

Two independent authors (MM and FM) assessed the language suitability and subject matter of each paper. Studies thereby selected were evaluated for their appropriateness for inclusion and quality of the method. The first author, year of publication, design, participants, country, assessment, instruments, and main findings are reported in **Table 1**.

**Table 1:**
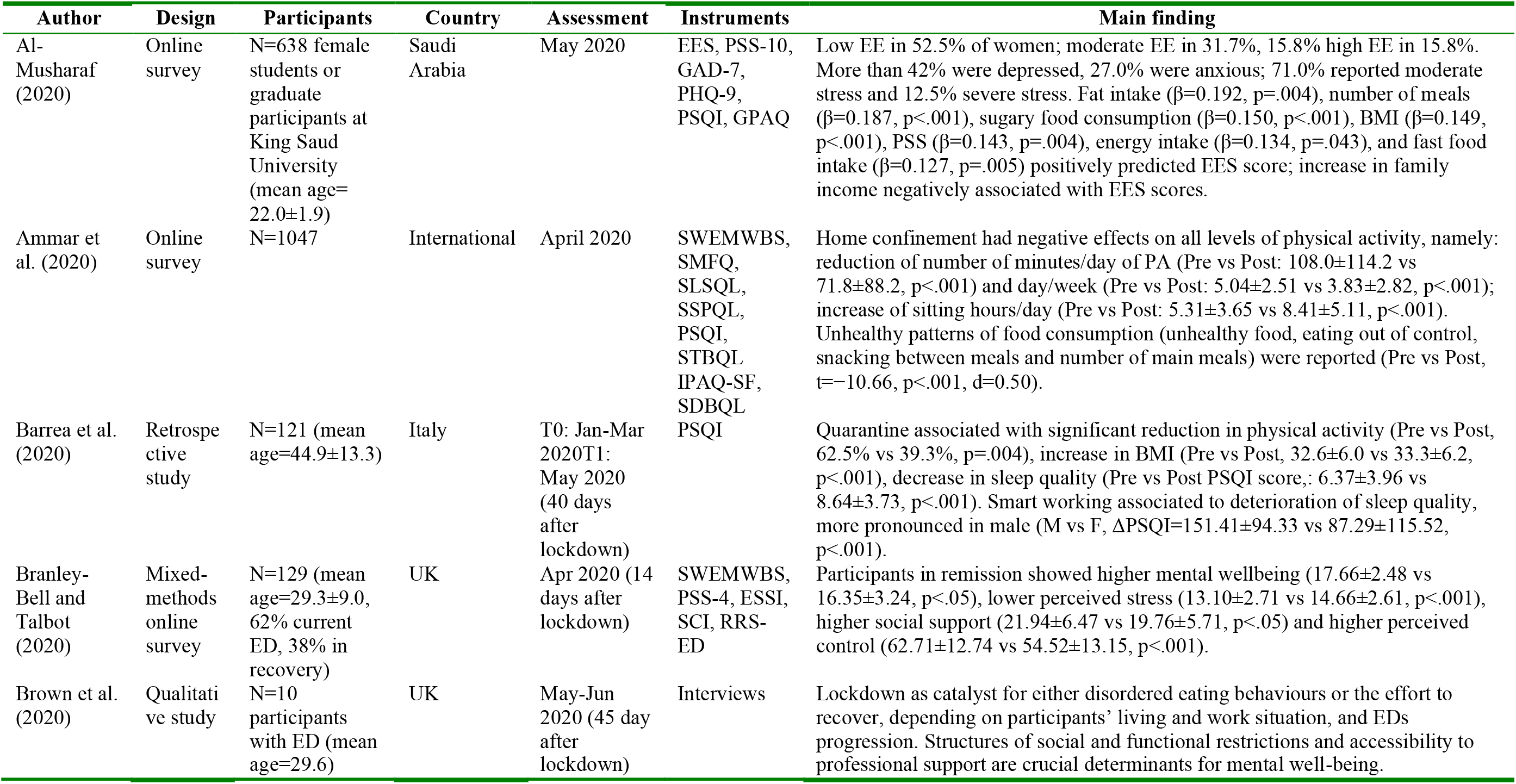
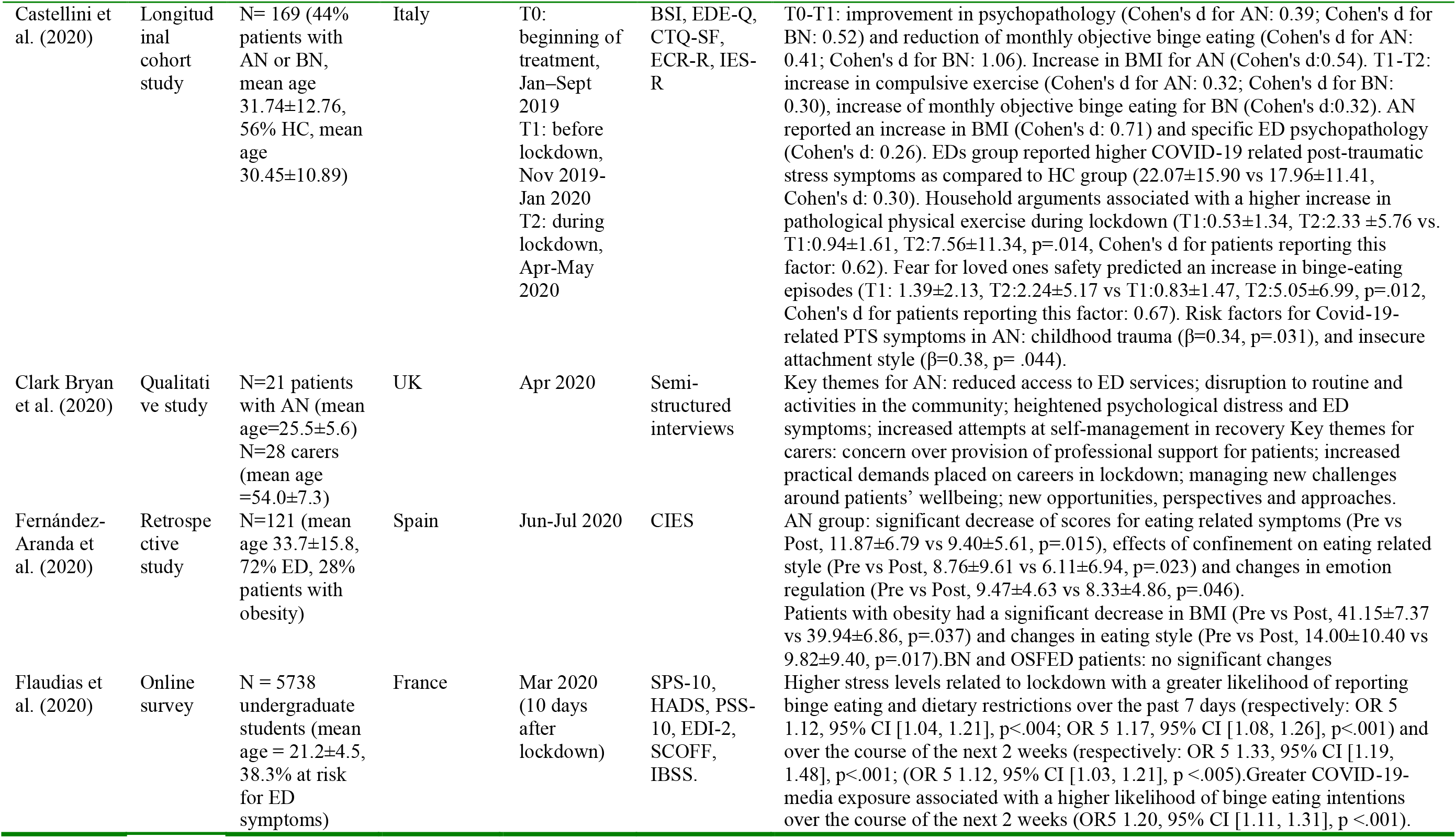
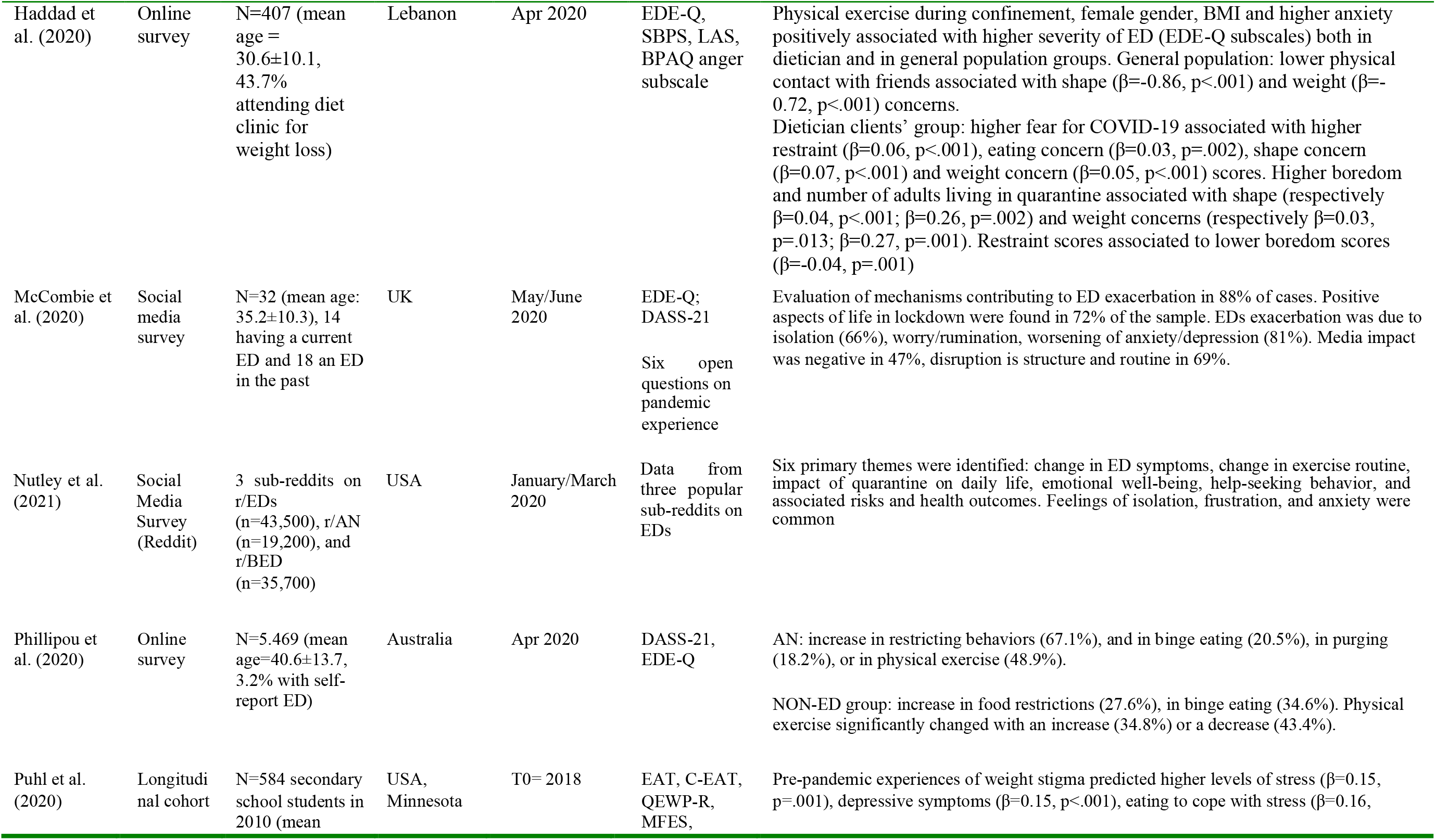
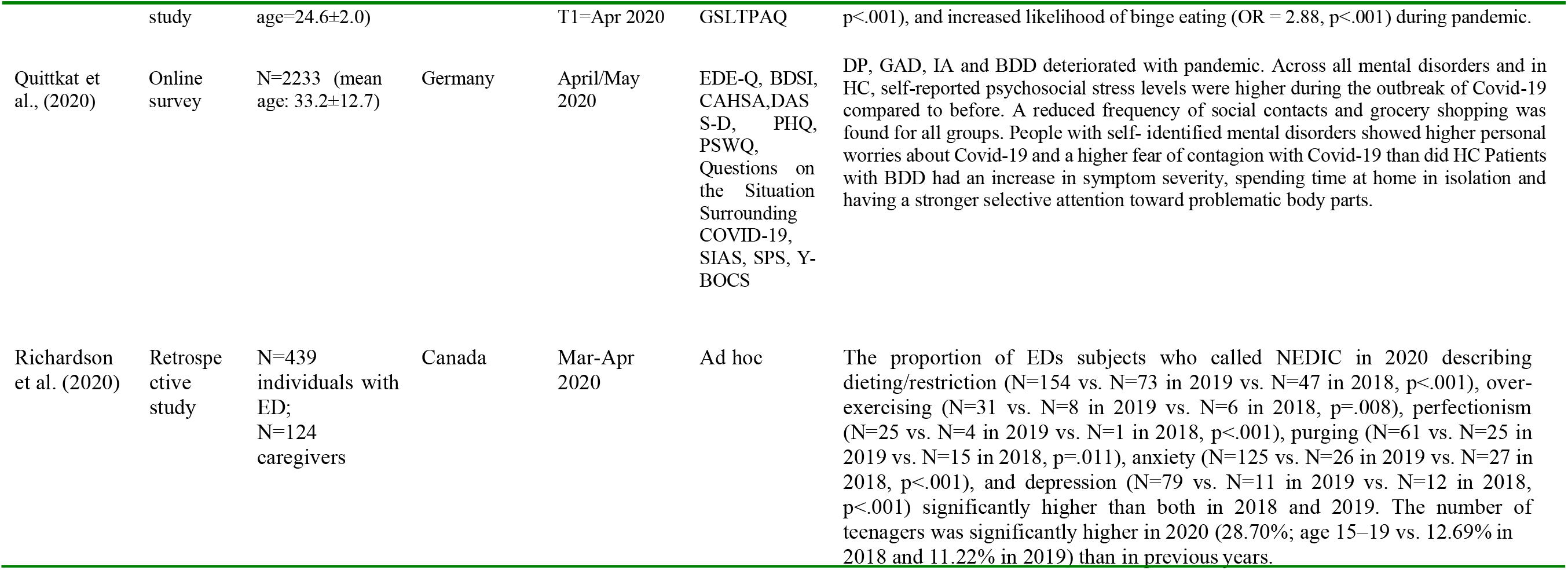
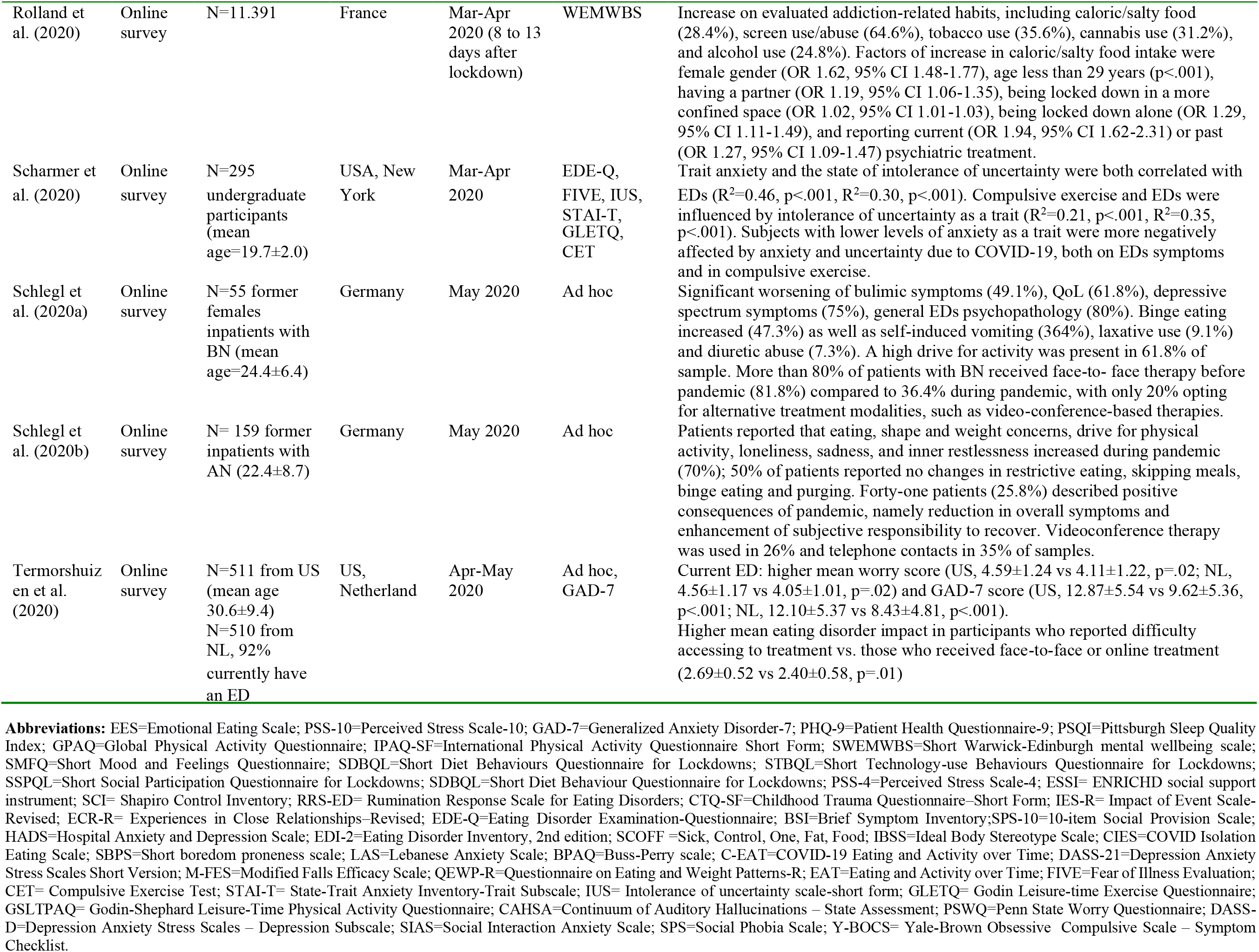
Studies on Eating Disorders Spectrum during COVID-19 pandemic.

### 2.6 Risk of bias in individual studies

Risk of bias of individual studies was not performed considering that no RCTs or systematic follow-up studies deriving from RCTs were found.

### 2.7 Data synthesis

A meta-analysis could not be performed because of the lack of homogeneity among the resulting studies. In particular, studies varied in terms of how improvements were measured. Hence, this systematic review is summarized in a narrative synthesis.

## 3 RESULTS

The systematic search identified 91 studies of which 21 papers were eligible and were included (**Table 1**). Nine (42.9%) out of the total papers evaluated the effect of pandemic and associated protective and risk factors in EDs patients, ten (47.6%) explored the prevalence of disturbed eating behaviours and risk factors for exacerbating EDs in the general population, and the remaining two (9.5%) were qualitative studies describing the impact of lockdown and quarantine on EDs patients. The majority of studies collected data through online platforms (online survey). We sub-divided the retrieved studies according to their main research areas, as follows: studies on EDs in the general population; COVID-19 and lockdown consequences in EDs patients; key themes for patients with EDs during pandemic.

### 3.1 Eating spectrum in the general population

During COVID-19 pandemic, several studies investigated the prevalence of disturbed eating behaviours and related risk factors for exacerbations of EDs in the general population.

Al-Musharaf and colleagues (2020) evaluated emotional eating (EE) defined as the tendency to overeat as a coping strategy for regulating and reducing negative emotions (Ganley, 1989) in female students or graduate participants at King Saud University. Their study aimed to assess the impact of COVID-19 pandemic on EE during the lockdown and to identify indicators and predictors. A total of 1037 participants consented to take part in the study and, out of these, 638 were eligible. Participants completed the online questionnaire that included the Arabic version of the ‘*Emotional Eating Scale*’ (EES) Saade et al., 2019), ‘*Perceived Stress Scale*’ (PSS) (Cohen, Kamarck, & Mermelstein, 1983), ‘*Generalized Anxiety Disorder-7*’ (GAD-7) (Spitzer, Kroenke, Williams, & Löwe, 2006), ‘*Patient Health Questionnaire-9*’ (PHQ-9) (Kroenke, Spitzer, & Williams, 2001), ‘*Pittsburgh Sleep Quality Index*’ (PSQI) (Buysse, Reynolds, Monk, Berman, & Kupfer, 1989) and ‘*Global Physical Activity Questionnaire*’ (GPAQ) (World Health Organization, 2005). Low EE was reported by 52.5% of women, while 31.7% moderate and 15.8% high stress. As expected, BMI was significantly greater in the high EE group than in moderate and low groups. A total of 42.8% of the women reported depressive symptoms, 27.0% anxiety, 71.0% moderate stress and 12.5% severe stress. Higher perceived stress, BMI, fat intake, number of meals per day, sugary food consumption and frequent fast food intake positively correlated with EE scores. No relationship was found between EE and anxiety, depression, physical activity and sleep quality. The authors speculated that, under the stressful circumstances of COVID-19 pandemic, EE might be interpreted as an adaptive mechanism for managing negative emotions. Surprisingly, both baseline low family income and increased family income during the pandemic were negatively associated with EE scores, however, this finding, albeit intriguing requires further investigations.

The study of Ammar and colleagues (2020) reported the preliminary data from an online survey collected in a sample of 1047 participants from Asia (36%), Africa (40%), Europe (21%) and others (3%). The study was part of a larger survey that assessed the ‘Effects of home Confinement on multiple Lifestyle Behaviours during the COVID-19 outbreak (ECLB-COVID19)’. Questions were presented for comparing ‘*before*’ vs. ‘*during*’ confinement conditions. The authors investigated changes in physical activity with the ‘*International Physical Activity Questionnaire Short Form*’ (IPAQ-SF) (Craig et al., 2003), and dietary behaviours during lockdown with a newly developed crisis-oriented questionnaire, the ‘*Short Diet Behaviour Questionnaire for Lockdowns’* (SDBQ-L) (Brach et al., 2020). The authors highlighted that participants increased the consumption of unhealthy food, snacking between meals, eating out of control, and the frequency of main meals. The total score of diet increased significantly during confinement. Moreover, the time spent for physical activity decreased under confinement condition (days/week, 24%; minutes/day, 33.5%) meanwhile sedentary behaviour, such as the daily sitting time, increased more than 28% (from 5 to 8 hours per day). As a result, home confinement during COVID-19 pandemic dramatically reduced physical activity and altered eating behaviours in an unhealthy way.

One retrospective study investigated the effects of quarantine on BMI and sleep quality in Italian adults (N=121) (Barrea et al., 2020). Data were collected at baseline in their obesity outpatient clinic and after 40 days of quarantine by telephone interview (T1). Physical activity, working modalities and sleep quality were assessed at T1. Sleep quality was evaluated with ‘*Pittsburgh Sleep Quality Index*’ (PSQI) (Buysse et al., 1989). All participants reported a reduction in physical activity during the quarantine. After the quarantine, a significant increase in BMI was observed, particularly in normal weight and obesity of grade I, and II. Meanwhile, sleep quality worsened, except for severe obesity (grade III). Smart working negatively influenced sleep quality in all participants, with a greater worsening effect in men.

The study of Flaudias and colleagues (2020) is an ancillary project drawn from a larger database. An online questionnaire was distributed over 2 days to French undergraduate students (N=5738), 10 days after the declaration of the lockdown. Participants were evaluated with ‘*10-item Social Provision Scale*’ (SPS-10) (Cutrona & Russell, 1983), ‘*Hospital Anxiety and Depression Scale*’ (HADS) (Zigmond & Snaith, 1983), ‘*Perceived Stress Scale*’ (PSS) (Cohen et al., 1983), body dissatisfaction and impulse regulation subscales of the ‘*Eating Disorder Inventory, 2nd edition*’ (EDI-2) (Garner, 1991), an EDs screening tool, ‘*Sick, Control, One, Fat, Food*’ (SCOFF) (Garcia et al., 2010), and the ‘*Ideal Body Stereotype Scale*’ (IBSS) (Stice & Agras, 1998). Variables such as lockdown-related stress, COVID-19-related stress and media exposure to COVID-19 were also collected. The increased stress related to the lockdown, social distancing and disruptions in daily routines, was associated with a greater likelihood of reporting binge eating and dietary restrictions during the past week, and with higher intentions to engage in such problematic eating behaviours over the next two weeks. Moreover, greater exposure to COVID-19-related media was associated with the intent to engage in binge eating. Meanwhile, a relationship between concerns related to COVID-19 pandemic itself, such as fear of contagion, and disturbed eating behaviours was not found. As expected, pre-existing eating concerns were identified as risk factors for the development of problematic eating behaviours during stressful situations, such as the COVID-19 pandemic and lockdown are. In summary, the authors highlighted a strong relationship between problematic eating behaviours and stress related to the lockdown. The measure of stress used in the study included social distancing and disruption of daily routines that might promote a wide range of negative effects and increase problematic eating behaviours.

Haddad and colleagues (2020) compared a group of people attending diet clinic for weight loss management (N=177), supposed to have more problematic eating behaviours, and a group from the general population (N=228) in Lebanon. The study aimed at evaluating the association between lockdown stressors and eating behaviours and to compare the two groups of participants in terms of vulnerability. Participants were evaluated regarding quarantine and confinement stressor, current fear of COVID-19, and by using the ‘*Lebanese anxiety scale*’ (LAS) (Hallit et al., 2020), ‘*Short boredom proneness scale’* (SBPS) (Struk, Carriere, Cheyne, & Danckert, 2017), ‘*Eating Disorder Examination-Questionnaire*’ (EDE-Q) (Fairburn & Beglin, 2008) and the anger subscale of the ‘*Buss-Perry scale*’ (Buss & Perry, 1992). The authors found that 44.8% of participants exhibited a high fear of COVID-19. Physical exercise during confinement, female gender, BMI and higher anxiety were positively associated with higher severity of EDs, amongst both dietician’s clients and the general population. In participants with a higher risk for developing problematic eating behaviours (dietician clients’ group), the higher EDs severity was associated with a greater fear of COVID-19. This result partially confirmed that stressful events might worsen disturbed eating behaviours, particularly in vulnerable population. In this study, the authors highlighted that social distancing was associated with shape and weight concerns only in the control group. Meanwhile, amongst dietician’s clients’, both a higher boredom and number of adults living in the same house during the quarantine were associated with higher shape and weight concern scores, while confirming the negative impact of functional restrictions and changes in daily routines on disturbed eating behaviours.

The study by Phillipou et al. (2020) explored changes in eating and exercise behaviors following the official announcement of COVID pandemic in Australia, comparing subjects with and without a history of EDs in a general population sample, aged >18 years. The main hypothesis of the study was that subjects with a self-reported history of EDs might report significant changes in their habits more than subjects without EDs, especially in four behaviors during the pandemic, namely: restricting, binge-eating, purging and physical exercise. The study was conducted with a series of 72 hours open anonymous online surveys, each month, for one year, and was part of a larger survey, named COLLATE (COVID-19 and you: mental health in Australia now survey). Of the original 8,014 individuals enrolled in the COLLATE survey, 5,469 completed the eating and exercise behaviors section. Subjects with a positive a history of EDs were 180, of whom 88 with AN, 23 with BN, 6 with BED, 4 with ED-NOS and 68 who did not specify the EDs subtype. Given the diagnostic distribution, the most interesting findings were from the AN subgroup. Indeed, these subjects noted significant change in their eating and exercise behaviors, with an increase in restricting behaviors (67.1%), in binge eating (20.5%) or in purging (18.2%). Half of the subjects (48.9%) reported increased physical exercise in order to achieve more control on body shape and caloric intake. The same variables investigated in the subjects with no EDs, did not reveal any changes in the amount of food restricted, or on binge eating or purging behaviors. Only the 27.6% of the no-EDs sample reported a greater level of food restriction than before COVID-19. Conversely, 34.6% reported an increase in binge eating spectrum behaviors. Only one-third of the sample (34.8%) showed an increase in physical exercise routine, whereas 43.4% reported a decrease e the remaining no change. To summarize, this study provided evidence regarding the worsening of eating spectrum signs, symptoms and behaviors during pandemic, mainly when a history of EDs was present. Interestingly, the most represented diagnostic group was AN, while BN and BED seems to be rare. It is also noteworthy that body dysmorphic disorder (BDD) was detected as comorbid with AN only in the 1.1% of the sample. These findings are surprising and maybe due to the self-report design of the survey. We could speculate that AN subjects might be more reliable and more precise in describing their habits than their body dissatisfaction, if compared with subjects with BN or BED. We could also hypothesize that the ‘*restricter’* habits are less related to a negative stigma than bingeing or purging, and might be more easily self-reported in an epidemiological survey.

Weight stigma has been considered as a relevant variable in a study conducted on a subsample derived from a longitudinal cohort study, the EAT 2010-2018, and labeled as COVID-19 Eating and Activity over Time Study (C-EAT) (Puhl, Lessard, Larson, Eisenberg, & Neumark-Stzainer, 2020). The main aim of C-EAT survey was to explore how weight-related health behaviors may change as a result of events related to COVID-19. More in detail, it was considered how high body weight subjects perceived the social stigma linked to obesity during pandemic, and how such stigma contributed to the adoption of unhealthy eating behaviors, and a lower physical activity. Individuals who experienced pre-pandemic weight stigma were compared with those who did not. In agreement with the original assumption, the majority of the overall sample reported that COVID-19 negatively affected mood and anxiety levels, raising stress, and exacerbating problems with food intake. The comparison between the two samples (stigma perceived before COVID *vs*. stigma perceived during COVID) revealed that pre-pandemic experiences of weight stigma predicted higher levels of stress, depressive symptoms, eating to cope with stress, and an increased likelihood of binge eating among young adults during the COVID-19 pandemic. Furthermore, no significant interactions based on gender were found, indicating that these adverse health outcomes associated with weight stigma were present regardless of gender identity. In summary, young adults who experienced weight stigma before COVID showed increased vulnerability for binge eating and psychological distress during the pandemic. No information was provided regarding other bio-psychosocial variables, such as the interpersonal environment of subjects or the role of attachment figures.

Changes in caloric food intake were also found in a web-based survey carried out in France during the first days of containment due to COVID-19, on more than 11.000 participants (Rolland et al., 2020). Apparently, respondents reported an increase on all evaluated addiction-related habits, including caloric/salty food (28.4%). However, use/abuse of caloric foods was under-represented, if compared with screen use/abuse (64.6%), or tobacco (35.6%) and cannabis use (31.2%), and more represented than alcohol use (24.8%). Not surprisingly, female gender, age less than 29 years, being locked down in a more confined space, being locked down alone, and reporting current or past psychiatric treatment were all factors of increased caloric/salty food intake.

Scharmer et al. (2020) investigated the potential relationships between COVID-19 pandemic, anxiety levels, subjective intolerance of uncertainty and the risk for EDs pathology/compulsive exercise, in asample of 295 undergraduate women in USA. Subjects were evaluated with the ‘*Eating Disorder Examination-Questionnaire*’ (EDE-Q)(Berg, Peterson, Frazier, & Crow, 2012; Fairburn & Beglin, 2008), the ‘*Fear of Illness Evaluation*’ (FIVE) (Ehrenreich-May, 2020), the ‘*State-Trait Anxiety Inventory-Trait Subscale*’ (STAI-T) (Barnes, Harp & Young, 2002), the ‘*Compulsive Exercise Test*’ (CET) (Taranis, Touyz, & Meyer, 2011), the ‘*Intolerance of uncertainty scale-short form*’ (IUS) (Nicholas Carleton, Sharpe, & Asmundson, 2007) and the ‘*Godin Leisure-time Exercise Questionnaire*’ (GLETQ) (Godin, 2011). They basically explored the effects of four different variables on EDs and compulsive exercise, namely the levels of anxiety correlated to COVID-19 (‘*anxiety as a state*’), the intolerance to uncertainty correlated to COVID-19 (‘*intolerance as a state*’), the ‘*anxiety levels*’ as a stable trait, and the ‘*levels of intolerance to uncertainty*’ as a trait. Their results showed that both the state anxiety and the state of intolerance of uncertainty were positively related to EDs, but not with compulsive exercise. By contrast, compulsive exercise and EDs were influenced by the intolerance of uncertainty as a trait. Moreover, subjects with lower levels of pre-existing COVID-19 anxiety (i.e., anxiety as a trait) were the most negatively affected by anxiety and uncertainty due to COVID-19, both on EDs symptoms and in compulsive exercise. In summary, the authors concluded that stable traits of anxiety and uncertainty were more relevant in worsening EDs and compulsive exercise than the current levels of the same subjective variables.

A new approach to the evaluation of disordered eating behaviors in the general population has been adopted by Nutley et al. (2021) that analyzed the experiences posted on the internet-based social media platform Reddit, by worldwide users, consisting in stories, web content ratings and discussion communities. They coded posts as belonging to categories used to construct specific themes. Six ‘*primary themes*’ emerged, namely: worsening in EDs symptoms (including, negative body image), change in exercise routine (also because exercise facilities were closed or inaccessible), impact of quarantine on daily life (change in routine/environment, food hoarding or shortages) emotional well-being, help-seeking behavior (willingness to recover, inability to receive treatments, advices’ requests from other Reddit users, seeking help), associated risks (including, substance use behaviors), and adverse health outcomes. Feelings of isolation, frustration, and anxiety were also described as common.

### 3.2 COVID-19 and lockdown consequences in patients with EDs

Branley-Bell & Talbot (2020) collected data from 80 subjects with current EDs and 49 in recovery with the aim of understand the impact of COVID-19 pandemic in people with experiences of ED. A mixed-methods online survey was developed. Participants completed open-ended questions related to the impact of lockdown and some validated questionnaires to assess mental well-being, ‘*Short Warwick-Edinburgh mental wellbeing scale*’ (SWEMWBS) (Tennant et al., 2007), perceived stress, ‘*Perceived Stress Scale*’ (PSS) (Cohen et al., 1983), social support, *‘ENRICHD social support instrument’* (ESSI)(Vaglio et al., 2004), perceived control, ‘*Shapiro Control Inventory*’ (SCI) (Shapiro, 1994) and rumination, ‘*Rumination Response Scale for Eating Disorders*’ (RRS-ED) (Cowdrey & Park, 2011). The majority of participants (86.7%) reported that ED symptoms had worsened as a result of the pandemic. Only 2 participants reported an improvement in symptoms. Participants in remission exhibited higher mental wellbeing, lower perceived stress, higher social support, and higher perceived control as compared with participants with a current ED.

The study of Fernández-Aranda and colleagues (2020) aimed at evaluating the impact of home-confinement, in terms of changes in eating behaviours and symptoms, in a sample of 87 EDs patients and 34 obese patients. EDs patients were recruited in centres representative of the public and private sectors of ED treatment services in Barcelona: 55 were suffering from AN, 18 from BN, and 14 from other specified feeding or eating disorders (OSFED), according to DSM-5 criteria. Obese patients were recruited at the Endocrinology Unit at the Clinic Hospital of Barcelona. The authors collected data with the COVID Isolation Eating Scale (CIES), a newly created questionnaire that investigated four different domains: circumstances during confinement, effects of confinement on EDs symptoms, behavioural and psychopathological impact of confinement and evaluation of online intervention, considering ‘before’ *vs*. ‘after confinement’. Psychometric properties of the CIES were analysed and factor analysis confirmed the rational structure of the CIES. In contrast with previous studies, the disordered eating improved in almost all participants, during confinement. The impact of COVID-19 pandemic was mixed and varied in the given diagnostic distribution. Surprisingly, AN patients reported a decrease in EDs symptoms and in emotion dysregulation; obese patients showed a significant decrease in BMI and in eating symptomatology; BN and OSFED patients exhibited no significant changes before and after confinement. Patients with OSFED deserved special attention: although changes between pre- and post-confinement did not reach the statistical significance, they reported the highest impairment in psychopathology. This finding highlighted the need to promptly identify vulnerable subjects that may be more sensitive to adverse events. Concerning the use of telemedicine, AN patients expressed the greatest dissatisfactions and difficulties with online treatments.

The longitudinal study of Castellini and colleagues (2020) aimed to test three main hypotheses: EDs patients might represent a more vulnerable population to the effects of COVID-19 pandemic; recovery process might be affected by the lockdown circumstances; finally, factors preceding pandemic might be associated with worsening of psychopathology during lockdown. Data were collected from 74 patients with a current DSM-5 diagnosis of AN or BN, attending an individual Enhanced Cognitive Behavioural Therapy (Fairburn, 2008) at an Outpatient Clinic for EDs. A group of 97 healthy controls (HC) was also enrolled. Both groups were evaluated before lockdown (T1) and during lockdown (T2), while AN and BN patients were also evaluated at the beginning of the treatment (T0). T0, T1 and T2 assessment included ‘*Brief Symptom Inventory*’, (BSI) (Derogatis, 1983) and ‘*Eating Disorder Examination Questionnaire*’, (EDE-Q) (Fairburn & Beglin, 2008). At T0 the ‘*Childhood Trauma Questionnaire-Short Form*’, (CTQ-SF)(Bernstein et al., 2003) and the ‘*Experiences in Close Relationships–Revised*’, (ECR-R) (Fraley, Waller, & Brennan, 2000) were also administered. Moreover, during lockdown (T2), the ‘*Impact of Event Scale-Revised*’ (IES-R) (Weiss & Marmar, 1997), adapted for COVID-19 pandemic, was administered. Not surprisingly, ED patients were more vulnerable to lockdown effects than HC, as confirmed by their increase in pathological eating behaviours (objective binge eating and compensatory physical exercise). Moreover, EDs group reported higher COVID-19 related post-traumatic stress symptoms, as compared with HC group. While comparing baseline to T1, EDs patients reported an improvement of general psychopathology and ED specific symptoms, objective binge eating and compensatory physical exercise. Moreover, BMI increased significantly in AN patients, while remained stable in BN patients. Comparisons between pre- and post-lockdown (T1-T2) revealed that EDs patients increased physical exercise during lockdown, despite the initial improvement. Some specific differences between diagnostic subgroups emerged: AN patients reported an improvement in BMI and specific EDs psychopathology, while BN patients exhibited an increase of objective binge eating. Despite ten full remissions and 19 partial remissions at T1, ten subjects reported relapse into BN at T2. No significant change was found in the HC group. Interestingly, the main factor of increase in pathological physical exercise was household arguments, while fear of safety for the loved one predicted a higher increase in binge eating episodes. Childhood trauma and insecure attachment style resulted to be factors for COVID-19-related PTS symptoms in AN patients. In summary, EDs patients, and particularly those suffering from BN were more vulnerable to the impact of lockdown. Indeed, lockdown interfered with recovery process in terms of relapses into pathological eating behaviours, such as compulsive physical exercise and exacerbations of binge eating. Results in AN patients appeared more controversial, as BMI and EDs psychopathology constantly improved, despite the exacerbation of compulsive exercise.

Changes in help-seeking behaviors from EDs subjects and their caregivers due to COVID pandemic was the focus of a Canadian study that retrospectively analyzed part of the information collected by the National Eating Information Centre (CEDIC), a national no-profit organization that, since 1985, operates toll-free helpline and instant chat service for patients with eating disorders (Richardson, Patton, Phillips, & Paslakis, 2020). The proportion of EDs patients who called the CEDIC describing dieting/restriction, over-exercising, perfectionism, purging, anxiety, and depression was significantly higher in 2020 than in both 2018 and 2019. The most relevant issues in 2020 were related to the difficulties in accessing treatments because of the pandemic, and to an urgent need for support, due to a worsening of symptoms, and a subjective feeling of losing control over eating symptoms and behaviors, with significant changes in food intake. Interestingly, there were a significantly higher number of teenagers who contacted NEDIC in 2020 compared to 2018 and 2019 (28.70%; age 15-19 vs. 12.69% in 2018 and 11.22% in 2019), maybe because teenagers were not attending school in person and had no regular access to their usual support networks and structures (guidance counselors, or teachers).

Schlegl and Colleagues (2020) published two papers based on an online survey for patients with EDs, interviewed during COVID-19 pandemic. The first study was carried out in a sample of 55 former female inpatients with BN (age range 17-46 years) (Schlegl, Meule, Favreau, & Voderholzer, 2020). They were evaluated by a self-developed questionnaire assessing psychological consequences of pandemic, such as the overall impact on EDs symptoms and on quality of life (QoL), the adverse effect on therapies, the incidence of new symptoms, worries regarding infections, relapses, food insecurity or job, general psychopathology, interpersonal conflicts and health care utilization. Enjoyable activities, virtual social contact with friends and mild physical exercises were also considered among the helpful strategies adopted by patients to face pandemic stressful events. In summary, results were discouraging: more than half of interviewed patients described a significant worsening of their bulimic symptoms and QoL. They also described a worsening of depressive spectrum signs and symptoms together with general psychopathology levels in around 80% of cases. Binge eating increased as well as self-induced vomiting, laxative use and diuretic abuse. A high drive for activity was present in around 60% of sample. Moreover, patients described difficulties with daily routine, especially for meals. The worsening of symptoms was also related to the interruption of face-to-face therapies in more than half of patients, with only 20% opting for alternative treatment modalities, such as video-conference-based therapies.

The second study was conducted on a sample of 159 former AN inpatients (age range 14-62 years), with the same modalities of that on BN patients (Schlegl, Maier, et al., 2020). In this case, results were less discouraging. Indeed, even if 70% of patients reported that eating, shape and weight concerns, drive for physical activity, loneliness, sadness, and inner restlessness increased during pandemic, there was also a sub-sample of 41 patients (25.8%) describing positive consequences, in terms of a reduction in overall symptoms and of an enhancement of subjective responsibility to recover. Moreover, videoconference therapy was used by 26% and telephone contacts by 35% of patients.

A two-site-online study in USA and the Netherlands recruited subjects with EDs (AN, BN, and BED) via social media, or through emails when they were already enrolled in previous surveys (Termorshuizen et al., 2020). In this study, variables such as exposure to virus, diagnosis in self or relatives, impact of COVID-19 on family members’ health or employment, and level of lockdown were collected together with the impact of pandemic on EDs, that was evaluated with a 4-point Likert-scale asking general questions on health subjective perception and specific questions on symptoms, such as binge eating. A 7-point Likert scale was also administered to explore worries about exposure and/or contracting the virus. Finally, a free text item inquired changes to EDs treatments, including frequency of treatment sessions and subjective perception of quality of treatments. Patients with AN described mainly fears about the availability of foods chosen for their ‘special’ meals. Moreover, they reported an exacerbation of food restriction. Patients with BN and BED increased binge and purge episodes. Positive effects of pandemic were described also in this survey, especially on both the interpersonal (greater connection with relatives) and the motivational side. All groups described difficulties in accessing their treatments.

Two studies reported inconclusive and countertrend results. The first one was an online survey conducted in UK on a very small sample of 32 subjects >17 years old with current or previous EDs, invited with posts on Twitter (McCombie, Austin, Dalton, Lawrence, & Schmidt, 2020). Participants were asked to describe their EDs status (current/partial recovery/full recovery), and to report on their illness duration. They were evaluated with the “*Eating Disorder Examination Questionnaire*” (EDE-Q), the “*Depression, Anxiety and Stress Scales-Version 21*” (DASS-21), and with a six open-ended questions interview, exploring the subjective experience of pandemic. EDs symptoms were described as exacerbated by pandemic, and by the associated changes in lifestyle in 88% of cases. Physical and psychological isolation, worry about the future, fear around a ‘*return to normal life*’, changes in daily routine were the main topics. However, the 72% of participants described also the positive effect of having more space and time for healing and self-care, and having less pressure to engage in social activities. The second study was an online survey conducted in Germany on a heterogeneous sample (n=2233), in which several mental disorders (including EDs) were assessed (Quittkat et al., 2020). Participants were recruited through university press releases, e-mail lists, flyer, social media, institutions for education in psychotherapy, outpatients departments, mental hospitals, self-help groups and assisted living departments; comparisons were made between individuals with and without mental disorders. According to this exploratory study, no changes in the level of EDs symptoms were found, and no trend toward a change in pathology.

### 3.3 Key themes for patients with EDs during pandemic

Three qualitative studies explored the impact of COVID-19 outbreak and associated lockdown measures on patients with self-reported EDs (Branley-Bell & Talbot, 2020; S. M. Brown et al., 2020) or patients with AN and their carers (Clark Bryan et al., 2020).

As a consequence of the intense media reports on COVID-19 8 (the so-called ‘*infodemic*’), levels of anxiety increased, with an exacerbation of fear of contamination and washing compulsions (Clark Bryan et al., 2020). Some patients revealed that fear of contamination concerned primarily the health of the elderly and more vulnerable loved ones rather than themselves (S. M. Brown et al., 2020).

The introduction of lockdown measures and social isolation was associated with negative mental health outcomes, sense of loneliness, tendency to become more focused on food, rumination over food intake, disordered eating behaviours, such as restrictions or binge eating, and compulsive physical exercise (Branley-Bell & Talbot, 2020; S. M. Brown et al., 2020; Clark Bryan et al., 2020).

Moreover, the imposed functional restrictions disrupted daily-established routines that needed to be adapted to the uncertainty of the continuously changing environment. Again, patients reported increased anxiety for the need to deconstruct rigid regimens and to adjust them to the current situation (S. M. Brown et al., 2020) and experienced reduced motivation for recovery (Branley-Bell & Talbot, 2020; Clark Bryan et al., 2020). This led to exacerbate EDs symptoms and to actively engage in disordered eating behaviours that provided a sense of control (Branley-Bell & Talbot, 2020).

Besides, the increased amount of time spent online harmed those suffering for EDs. The enhance exposure to triggering contents, such as messages promoting in-house physical exercise or food recipes, and the easy access to extreme content (such as pro-AN content) was detrimental for most participants (Branley-Bell & Talbot, 2020). Only social support perceived through social media (WhatsApp, etc) represented a resource for reducing the lockdown sense of isolation (Branley-Bell & Talbot, 2020).

Brown and colleagues (2020) pointed out that lockdown measures might also have a positive effect and lead to recovery, depending on the participants’ working and living situation (alone vs. with family or partner or friends), and EDs progression. Some participants reported increased self-responsibility, intentionality in planning their actions, and increased attempts for a self-management that improved their EDs behaviours.

All studies highlighted the role of the accessibility to professional support as a crucial determinant of mental wellbeing: patients complained about discrepancies in service provision, premature discharge and suspension of treatments that increased anxiety and feelings of being abandoned. (Branley-Bell & Talbot, 2020; S. M. Brown et al., 2020; Clark Bryan et al., 2020). Although the continuation of treatments through online delivery was described as a positive factor, this was not seen as a ‘valuable alternative’ but just as the ‘only possibility’ (Branley-Bell & Talbot, 2020).

## 4 DISCUSSION

COVID-19 pandemic significantly modified daily life, interpersonal relationships, and individual perception of the future, thus promoting anxiety, stress, and uncertainty (Conversano et al., 2020). From a psychological perspective, pandemic provoked negative effects on mental wellbeing in two ways: first, patients already affected by EDs might experience a worsening of their clinical conditions and QoL; second, psychiatric disorders, such as EDs, might be triggered in predisposed subjects who had a diathesis for a specific disorder by the exceptional conditions deriving from COVID-19 related stress. Therefore, studies in EDs focused their attention on both areas. We list herein the most relevant issues collected from selected observations:

- *Food access limitation* is considered by the majority of studies as one of the most relevant risk factors for both triggering and exacerbating EDs during COVID-19 pandemic. Subjects who previously had never experienced ‘*food insecurity*’ described, for the first time, an intense fear of having limited access to shops, or of finding empty shelves, long lines, and unavailability of specific foods considered essential to their diets. Patients already suffering for AN-R, or subjects with avoidant/restrictive food intake spectrum habits not fulfilling diagnostic criteria for an EDs, reported a disturbing experience of reduced access to their ‘*comfortable types and brand of foods*’. As a consequence, they skipped meals or exacerbated dietary restraint in order to ration the ‘*essential types of food they needed*’. Equally, food insecurity may trigger food hoarding and overconsumption with compensatory behaviors in subjects with AN-BP, BN or BED. Moreover, even in subjects with no diagnosis of EDs, food insecurity may increase ideational polarization on meals, thus raising the risk of binge eating, and the potential consequences in terms of body image changes.
- *Changes in physical activities routines*: Patients with AN experienced a worsening of their preoccupation on weight and body shape due to the difficulties in continuing their compensatory behaviors, including compulsive physical exercise during pandemic. However, we would propose a different interpretation of this phenomenon described by several studies, as we are of the opinion that one of the most relevant issues during pandemic is the overall change in daily routines, including, although not exclusively, the limitations of physical activities. For example, the ‘*social zeitgeber*’ hypothesis posits that unstable or disrupted daily routines might lead to circadian rhythm instability and, in vulnerable individuals, to mood instability, sleep disorders and dietary disruptions (Frank et al., 2005, 2008). According to this model, psychosocial factors would interact with chrono-biological mechanisms to create and/or facilitate a pathway towards psychopathology (Palagini et al., 2021).
- *Restrictions or difficulties in healthcare access:* This factor was obviously more relevant for EDs patients than for general population, considering that they might have unstable physical conditions requiring a frequent medical monitoring and lab tests. However, in several countries, the usual medical services have been converted to services exclusively dedicated to COVID patients, thus limiting the access for other medical reasons, and raising the subjective sense of uncertainty for the overall population (Marazziti et al., 2020). Telemedicine check-ups have been proposed as an alternative approach, but it is obvious that the monitor of weight changes, or vital signs is limited during a long-distance follow-up.
- *The role of anxiety levels:* The COVID pandemic represented and still represents an unpredictable general health threat, leading to a diffuse sense of uncertainty and a rising in anxiety levels. Several studies already highlighted how EDs patients might show higher levels of intolerance for uncertainty than general population (M. Brown et al., 2017). Moreover, as already pointed out, routines changes that could lead to body weight modifications may exacerbate additional concerns in EDs patients. Therefore, EDs stereotyped behaviors play a role in controlling anxiety levels and in managing negative emotions (Kesby, Maguire, Brownlow, & Grisham, 2017); both generalized uncertainty and anxiety associated with pandemic, and weight and shape specific uncertainty or changes, contributed to increase subjective discomfort. However, according to our observations, less importance has been given to the cognitive and affective sides of such discomfort, related to the interpersonal deficits deriving from pandemic, and on changes in interpersonal relationships, even if it is well-known that EDs can be triggered or worsened by interpersonal difficulties and/or disruptions. Binge eating and dietary restraint tend to occur in the context of (or are exacerbated by) adverse interpersonal events (Miniati, Callari, Maglio, & Calugi, 2018). During COVID-19 pandemic, characterized by protracted lockdowns and induced interpersonal deficits, EDs patients might have experienced a worsening of their symptoms as a direct consequence of deprivation in relationships with significant ones.
- *Social isolation and home confinement:* COVID-19 restrictions provoked a significant impact on social interactions. EDs patients, who were home-confined with family members, experienced feelings of scrutiny and pressure to recover one the one side, meanwhile conflicts and difficulties in relationships with ‘*no way out*’, acted as a maintenance factor for EDs symptoms. As other authors pointed out (Hillege, Beale, & McMaster, 2006), family relationships influenced the onset and maintenance of EDs symptoms, but also EDs might influence family dynamics, thus creating a negative feedback loop. For those who were living alone, the main challenge was to cope with the feeling of loneliness contributing to and fueling ED symptoms and vice-versa (Levine, 2012). As a consequence of the fact that nobody would see and recognize the loss of weight and/or physical deterioration, EDs patients easily increased compulsive pathological behaviors. Furthermore, some important elements of the recovery process, such as social eating, external distractions or physical connection with people, were replaced by rumination on food or physical activities and then caused relapses. Given that, although home confinement was highly commendable and necessary to limit the spread of the pandemic, the specific characteristics of EDs patients should be taken into account in order to plan specific interventions.
- *The problem of EDs in adolescents and young adults*: According to the current literature, no special attention has been devoted to the differences between adolescents/young adults and the rest of the populations, despite researches conducted before pandemic already demonstrated that approximately 85% of adolescents were unhappy with their body (Ricciardelli & McCabe, 2001), and that 50-60% of adolescents and 25% of emerging adults reported ‘*disturbed eating behaviors’* (e.g., fasting, taking diet pills or laxatives, vomiting, binge-eating) (Croll, Neumark-Sztainer, Story, & Ireland, 2002; Quick & Byrd-Bredbenner, 2013). Moreover, it is well known that maladaptive eating behaviors can be associated with several indicators of psychological distress, such as depressive symptoms or low self-esteem, independently from pandemic (Liechty & Lee, 2013), and that they represent major risk factors for the development of EDs (Neumark-Sztainer et al., 2006; Stice & Whitenton, 2002). Therefore, we were wondering on the ‘*specific*’role of COVID-19 pandemic, and on the relationships between new potential stressors and the worsening/triggering of EDs in adolescents/young adults. Even in this case, we found the same three main observations: concerns about access to and affordability of food, worries regarding infection, food insecurity, and, finally, limited access to psychological treatments, but this last factor seems to be of minor impact.

### 4.1 Study limitations

Across studies, the main weaknesses were due to limited assessment of pre-lockdown EDs symptoms, and to the absence of comparison groups. Moreover, in some studies, the EDs diagnoses were self-reported and specific information on disorders was not collected. Participants were mostly recruited online, via social media, resulting in samples being biased towards those who were familiar with the use of technology. The discrepancy in data collection points (early stage of lockdown vs. late stage) made difficult to compare the main findings.

## 5 CONCLUSION

During COVID-19 pandemic, medical practice should seek to maximize the access to treatments, while maintaining social distancing. It is essential that EDs patients remain in touch with their professional supports, to ensure that their health status is monitored. There is an urgent need to better understand the efficacy of the online delivery of treatments for EDs and to improve both the accessibility and acceptability (Barney, Buckelew, Mesheriakova, & Raymond-Flesch, 2020). We notice some criticisms of online surveys and long-distance management of EDs patients, such as limited accessibility, mainly because of technical issues (software, technical support etc.), difficulties experienced by clinicians in conducting online visits, the need for improvement in online software for the vital signs’ check, and privacy issues, especially for the youngest patients who are living with their relatives. Even with these limitations, telemedicine seems to be the most easy way in the mid-term for the management of EDs patients during pandemic, considering that we still do not know as yet how long pandemic will last, and that a number of ED patients may have physical complications of their psychiatric illness that raise risks in case of COVID-19 infection.

## Data Availability

Not applicable

## Authorship

All authors gave their substantial contributions to conception and design, data acquisition, data analysis, and interpretation. All authors gave contributions in drafting the article or critically revising it for important intellectual content, and gave their final approval of the version to be published. All authors agree to be accountable for all aspects of the work in ensuring that questions related to the accuracy or integrity of the work are appropriately investigated and resolved.

## Acknowledgements

None.

## Data Availability Statement

Not applicable

## Data deposition

not applicable

## REFERENCES

Al-Musharaf, S. (2020). Prevalence awend predictors of emotional eating among healthy young saudi women during the COVID-19 pandemic. Nutrients, 12(10), 1–17. https://doi.org/10.3390/nu12102923

Ammar, A., Brach, M., Trabelsi, K., Chtourou, H., Boukhris, O., Masmoudi, L., … Hoekelmann, A. (2020). Effects of COVID-19 home confinement on eating behaviour and physical activity: Results of the ECLB-COVID19 international online survey. Nutrients, 12(6). https://doi.org/10.3390/nu12061583

Barney, A., Buckelew, S., Mesheriakova, V., & Raymond-Flesch, M. (2020). The COVID-19 Pandemic and Rapid Implementation of Adolescent and Young Adult Telemedicine: Challenges and Opportunities for Innovation. Journal of Adolescent Health, 67(2), 164–171. https://doi.org/10.1016/j.jadohealth.2020.05.006

Barrea, L., Pugliese, G., Framondi, L., Di Matteo, R., Laudisio, D., Savastano, S., … Muscogiuri, G. (2020). Does Sars-Cov-2 threaten our dreams? Effect of quarantine on sleep quality and body mass index. Journal of Translational Medicine, 18(1), 318. https://doi.org/10.1186/s12967-020-02465-y

Berg, K. C., Peterson, C. B., Frazier, P., & Crow, S. J. (2012). Psychometric evaluation of the eating disorder examination and eating disorder examination-questionnaire: A systematic review of the literature. International Journal of Eating Disorders, 45(3), 428–438. https://doi.org/10.1002/eat.20931

Bernstein, D. P., Stein, J. A., Newcomb, M. D., Walker, E., Pogge, D., Ahluvalia, T., … Zule, W. (2003). Development and validation of a brief screening version of the Childhood Trauma Questionnaire. Child Abuse and Neglect, 27(2), 169–190. https://doi.org/10.1016/S0145-2134(02)00541-0

Brach, M., Trabelsi, K., Chtourou, H., Boukhris, O., Masmoudi, L., Bouaziz, B., … Hoekelmann, A. (2020). Effects of COVID-19 home confinement on physical activity and eating behaviour Preliminary results of the ECLB-COVID19 international online-survey. MedRxiv, 2020.05.04.20072447. https://doi.org/10.1101/2020.05.04.20072447

Branley-Bell, D., & Talbot, C. V. (2020). Exploring the impact of the COVID-19 pandemic and UK lockdown on individuals with experience of eating disorders. Journal of Eating Disorders, 8(1), 1–12. https://doi.org/10.1186/s40337-020-00319-y

Brown, M., Robinson, L., Campione, G. C., Wuensch, K., Hildebrandt, T., & Micali, N. (2017, September 1). Intolerance of Uncertainty in Eating Disorders: A Systematic Review and Meta-Analysis. European Eating Disorders Review, Vol. 25, pp. 329–343. https://doi.org/10.1002/erv.2523

Brown, S. M., Opitz, M. C., Peebles, A. I., Sharpe, H., Duffy, F., & Newman, E. (2020). A qualitative exploration of the impact of COVID-19 on individuals with eating disorders in the UK. Appetite, 156. https://doi.org/10.1016/j.appet.2020.104977

Buss, A. H., & Perry, M. (1992). The Aggression Questionnaire. Journal of Personality and Social Psychology, 63(3), 452–459. https://doi.org/10.1037//0022-3514.63.3.452

Buysse, D. J., Reynolds, C. F., Monk, T. H., Berman, S. R., & Kupfer, D. J. (1989). The Pittsburgh sleep quality index: A new instrument for psychiatric practice and research. Psychiatry Research, 28(2), 193–213. https://doi.org/10.1016/0165-1781(89)90047-4

Castellini, G., Cassioli, E., Rossi, E., Innocenti, M., Gironi, V., Sanfilippo, G., … Ricca, V. (2020). The impact of COVID-19 epidemic on eating disorders: A longitudinal observation of pre versus post psychopathological features in a sample of patients with eating disorders and a group of healthy controls. International Journal of Eating Disorders, (May), 1–8. https://doi.org/10.1002/eat.23368

Clark Bryan, D., Macdonald, P., Ambwani, S., Cardi, V., Rowlands, K., Willmott, D., & Treasure, J. (2020). Exploring the ways in which COVID-19 and lockdown has affected the lives of adult patients with anorexia nervosa and their carers. European Eating Disorders Review, (June), 1–10. https://doi.org/10.1002/erv.2762

Cohen, S., Kamarck, T., & Mermelstein, R. (1983). A global measure of perceived stress. Journal of Health and Social Behavior, 24(4), 385–396. https://doi.org/10.2307/2136404

Conversano, C., Marchi, L., & Miniati, M. (2020). Psychological distress among healthcare professionals involved in the COVID-19 emergency: Vulnerability and resilience factors. Clinical Neuropsychiatry, 17(2), 94–96. https://doi.org/10.36131/CN20200212

Cowdrey, F. A., & Park, R. J. (2011). Assessing rumination in eating disorders: Principal component analysis of a minimally modified ruminative response scale. Eating Behaviors, 12(4), 321–324. https://doi.org/10.1016/j.eatbeh.2011.08.001

Craig, C. L., Marshall, A. L., Sjöström, M., Bauman, A. E., Booth, M. L., Ainsworth, B. E., … Oja, P. (2003). International physical activity questionnaire: 12-Country reliability and validity. Medicine and Science in Sports and Exercise, 35(8), 1381–1395. https://doi.org/10.1249/01.MSS.0000078924.61453.FB

Croll, J., Neumark-Sztainer, D., Story, M., & Ireland, M. (2002). Prevalence and risk and protective factors related to disordered eating behaviors among adolescents: Relationship to gender and ethnicity. Journal of Adolescent Health, 31(2), 166–175. https://doi.org/10.1016/S1054-139X(02)00368-3

Cutrona, C., & Russell, D. W. (1983). The Provisions of Social Relationships and Adaptation to Stress. In Advances in Personal Relationships (pp. 37–67).

Derogatis, L. R. (1983). The Brief Symptom Inventory: An Introductory Report. Psychological Medicine, 13(3), 595–605. https://doi.org/10.1017/S0033291700048017

Fairburn, C. G. (2008). Cognitive behavior therapy and eating disorders. In Cognitive behavior therapy and eating disorders. New York, NY, US: Guilford Press.

Fairburn, C. G., & Beglin, S. (2008). EATING DISORDER EXAMINATION QUESTIONNAIRE (EDE-Q). Guilford Press, Cognitive.

Fernández-Aranda, F., Munguía, L., Mestre-Bach, G., Steward, T., Etxandi, M., Baenas, I.,…Jiménez-Murcia, S. (2020). COVID Isolation Eating Scale (CIES): Analysis of the impact of confinement in eating disorders and obesity—A collaborative international study. European Eating Disorders Review. https://doi.org/10.1002/erv.2784

Flaudias, V., Iceta, S., Zerhouni, O., Rodgers, R. F., Billieux, J., Llorca, P.-M., … Guillaume, S. (2020). COVID-19 pandemic lockdown and problematic eating behaviors in a student population. Journal of Behavioral Addictions, 9(3), 826–835. https://doi.org/10.1556/2006.2020.00053

Fraley, R., Waller, N., & Brennan, K. (2000). An item response theory analysis of self-report measures of adult attachment. Journal of Personality and Social Psychology, 78(2), 350–365. https://doi.org/10.1037//0022-3514.78.2.350

Frank, E., Kupfer, D. J., Thase, M. E., Mallinger, A. G., Swartz, H. A., Fagiolini, A. M., … Monk, T. (2005). Two-year outcomes for interpersonal and social rhythm therapy in individuals with bipolar I disorder. Archives of General Psychiatry, 62(9), 996–1004. https://doi.org/10.1001/archpsyc.62.9.996

Frank, E., Soreca, I., Swartz, H. A., Fagiolini, A. M., Mallinger, A. G., Thase, M. E., … Kupfer, D. J. (2008). The role of interpersonal and social rhythm therapy in improving occupational functioning in patients with bipolar I disorder. American Journal of Psychiatry, 165(12), 1559–1565. https://doi.org/10.1176/appi.ajp.2008.07121953

Ganley, R. M. (1989). Emotion and eating in obesity: A review of the literature. International Journal of Eating Disorders, 8(3), 343–361. https://doi.org/10.1002/1098-108X(198905)8:3<343::AID-EAT2260080310>3.0.CO;2-C

Garcia, F. D., Grigioni, S., Chelali, S., Meyrignac, G., Thibaut, F., & Dechelotte, P. (2010). Validation of the French version of SCOFF questionnaire for screening of eating disorders among adults. World Journal of Biological Psychiatry, 11(7), 888–893. https://doi.org/10.3109/15622975.2010.483251

Garner, D. (1991). Eating disorder inventory-2?: professional manual. Odessa Fla. (P.O. Box 998 Odessa 33556): Psychological Assessment Resources.

Godin, G. (2011). The Godin-Shephard Leisure-Time Physical Activity Questionnaire. Health and Fitness Journal of Canada, 4, 18–22.

Haddad, C., Zakhour, M., Bou Kheir, M., Haddad, R., Al Hachach, M., Sacre, H., & Salameh, P. (2020). Association between eating behavior and quarantine/confinement stressors during the coronavirus disease 2019 outbreak. Journal of Eating Disorders, 8(1). https://doi.org/10.1186/s40337-020-00317-0

Hallit, S., Obeid, S., Haddad, C., Hallit, R., Akel, M., Haddad, G., … Salameh, P. (2020). Construction of the Lebanese Anxiety Scale (LAS-10): a new scale to assess anxiety in adult patients. International Journal of Psychiatry in Clinical Practice, 24(3), 270–277. https://doi.org/10.1080/13651501.2020.1744662

Hensley. (2020). Why the coronavirus pandemic is triggering those with eating disorders - National | Globalnews.ca. Retrieved January 8, 2021, from https://globalnews.ca/news/6735525/eating-disorder-coronavirus/

Hillege, S., Beale, B., & McMaster, R. (2006). Impact of eating disorders on family life: Individual parents’ stories. Journal of Clinical Nursing, 15(8), 1016–1022. https://doi.org/10.1111/j.1365-2702.2006.01367.x

Kesby, A., Maguire, S., Brownlow, R., & Grisham, J. R. (2017, August 1). Intolerance of Uncertainty in eating disorders: An update on the field. Clinical Psychology Review, Vol. 56, pp. 94–105. https://doi.org/10.1016/j.cpr.2017.07.002

Kroenke, K., Spitzer, R. L., & Williams, J. B. W. (2001). The PHQ-9: Validity of a brief depression severity measure. Journal of General Internal Medicine, 16(9), 606–613. https://doi.org/10.1046/j.1525-1497.2001.016009606.x

Levine, M. P. (2012). Loneliness and eating disorders. Journal of Psychology: Interdisciplinary and Applied, 146(1–2), 243–257. https://doi.org/10.1080/00223980.2011.606435

Liechty, J. M., & Lee, M.-J. (2013). Longitudinal predictors of dieting and disordered eating among young adults in the U.S. International Journal of Eating Disorders, 46(8), 790–800. https://doi.org/10.1002/eat.22174

Marazziti, D., Pozza, A., Di Giuseppe, M., & Conversano, C. (2020). The psychosocial impact of COVID-19 pandemic in Italy: A lesson for mental health prevention in the first severely hit European country. Psychological Trauma: Theory, Research, Practice, and Policy, 12(5), 531–533. https://doi.org/10.1037/tra0000687

McCombie, C., Austin, A., Dalton, B., Lawrence, V., & Schmidt, U. (2020). “Now It’s Just Old Habits and Misery”–Understanding the Impact of the Covid-19 Pandemic on People With Current or Life-Time Eating Disorders: A Qualitative Study. Frontiers in Psychiatry, 11, 589225. https://doi.org/10.3389/fpsyt.2020.589225

McMenemy. (2020). Coronavirus and eating disorders: “I feel selfish buying food” - BBC News. Retrieved January 8, 2021, from https://www.bbc.com/news/uk-england-51962964

Miniati, M., Callari, A., Maglio, A., & Calugi, S. (2018). Interpersonal psychotherapy for eating disorders: Current perspectives. Psychology Research and Behavior Management, Vol. 11, pp. 353–369. https://doi.org/10.2147/PRBM.S120584

Miniati, M., & Marazziti, D. (2019, April 1). Psychopharmacological options for adult patients with anorexia nervosa: The patients’ and carers’ perspectives integrated by the spectrum model. CNS Spectrums, Vol. 24, pp. 225–226. https://doi.org/10.1017/S1092852917000700

Moher, D., Liberati, A., Tetzlaff, J., & Altman, D. G. (2009). Preferred Reporting Items for Systematic Reviews and Meta-Analyses: The PRISMA Statement. PLoS Medicine, 6(7), e1000097. https://doi.org/10.1371/journal.pmed.1000097

Neumark-Sztainer, D., Wall, M., Guo, J., Story, M., Haines, J., & Eisenberg, M. (2006). Obesity, disordered eating, and eating disorders in a longitudinal study of adolescents: How do dieters fare 5 years later? Journal of the American Dietetic Association, 106(4), 559–568. https://doi.org/10.1016/j.jada.2006.01.003

Nicholas Carleton, R., Sharpe, D., & Asmundson, G. J. G. (2007). Anxiety sensitivity and intolerance of uncertainty: Requisites of the fundamental fears? Behaviour Research and Therapy, 45(10), 2307–2316. https://doi.org/10.1016/j.brat.2007.04.006

Nutley, S. K., Falise, A. M., Henderson, R., Apostolou, V., Mathews, C. A., & Striley, C. W. (2021). Impact of the COVID-19 Pandemic on Disordered Eating Behavior: Qualitative Analysis of Social Media Posts. JMIR Mental Health, 8(1), e26011. https://doi.org/10.2196/26011

Orrù, G., Marzetti, F., Conversano, C., Vagheggini, G., Miccoli, M., Ciacchini, R., … Gemignani, A. (2021). Secondary Traumatic Stress and Burnout in Healthcare Workers during COVID-19 Outbreak. International Journal of Environmental Research and Public Health, 18(1), 337. https://doi.org/10.3390/ijerph18010337

Phillipou, A., Meyer, D., Neill, E., Tan, E. J., Toh, W. L., Van Rheenen, T. E., & Rossell, S. L. (2020). Eating and exercise behaviors in eating disorders and the general population during the COVID-19 pandemic in Australia: Initial results from the COLLATE project. International Journal of Eating Disorders, 53(7), 1158–1165. https://doi.org/10.1002/eat.23317

Poli, A., Gemignani, A., & Conversano, C. (2020). The psychological impact of sars-cov-2 quarantine: Observations through the lens of the polyvagal theory. Clinical Neuropsychiatry, 17(2), 112–114. https://doi.org/10.36131/CN20200216

Puhl, R. M., Lessard, L. M., Larson, N., Eisenberg, M. E., & Neumark-Stzainer, D. (2020). Weight Stigma as a Predictor of Distress and Maladaptive Eating Behaviors During COVID-19: Longitudinal Findings From the EAT Study. Annals of Behavioral Medicine?: A Publication of the Society of Behavioral Medicine, 54(10), 738–746. https://doi.org/10.1093/abm/kaaa077

Quick, V. M., & Byrd-Bredbenner, C. (2013). Disturbed eating behaviours and associated psychographic characteristics of college students. Journal of Human Nutrition and Dietetics, 26(SUPPL.1), 53–63. https://doi.org/10.1111/jhn.12060

Quittkat, H. L., Düsing, R., Holtmann, F. J., Buhlmann, U., Svaldi, J., & Vocks, S. (2020). Perceived Impact of Covid-19 Across Different Mental Disorders: A Study on Disorder-Specific Symptoms, Psychosocial Stress and Behavior. Frontiers in Psychology, 11, 3256. https://doi.org/10.3389/fpsyg.2020.586246

Ricciardelli, L. A., & McCabe, M. P. (2001). Children’s body image concerns and eating disturbance: A review of the literature. Clinical Psychology Review, 21(3), 325–344. https://doi.org/10.1016/S0272-7358(99)00051-3

Richardson, C., Patton, M., Phillips, S., & Paslakis, G. (2020). The impact of the COVID-19 pandemic on help-seeking behaviors in individuals suffering from eating disorders and their caregivers. General Hospital Psychiatry, 67, 136–140. https://doi.org/10.1016/j.genhosppsych.2020.10.006

Rolland, B., Haesebaert, F., Zante, E., Benyamina, A., Haesebaert, J., & Franck, N. (2020). Global Changes and Factors of Increase in Caloric/Salty Food Intake, Screen Use, and Substance Use During the Early COVID-19 Containment Phase in the General Population in France: Survey Study. JMIR Public Health and Surveillance, 6(3), e19630. https://doi.org/10.2196/19630

Saade, S., Hallit, S., Haddad, C., Hallit, R., Akel, M., Honein, K., … Obeid, S. (2019). Factors associated with restrained eating and validation of the Arabic version of the restrained eating scale among an adult representative sample of the Lebanese population: A cross-sectional study. Journal of Eating Disorders, 7(1), 24. https://doi.org/10.1186/s40337-019-0254-2

Scharmer, C., Martinez, K., Gorrell, S., Reilly, E. E., Donahue, J. M., & Anderson, D. A. (2020). Eating disorder pathology and compulsive exercise during the COVID-19 public health emergency: Examining risk associated with COVID-19 anxiety and intolerance of uncertainty. International Journal of Eating Disorders, (July), 1–6. https://doi.org/10.1002/eat.23395

Schlegl, S., Maier, J., Meule, A., & Voderholzer, U. (2020). Eating disorders in times of the COVID-19 pandemic—Results from an online survey of patients with anorexia nervosa. International Journal of Eating Disorders. https://doi.org/10.1002/eat.23374

Schlegl, S., Meule, A., Favreau, M., & Voderholzer, U. (2020). Bulimia nervosa in times of the COVID-19 pandemic—Results from an online survey of former inpatients. European Eating Disorders Review. https://doi.org/10.1002/erv.2773

Shapiro, D. H. (1994). Shapiro Control Inventory (SCI) Manual | Control Research. Retrieved from http://controlresearch.net/shapiro-control-inventory-manual.html

Spitzer, R. L., Kroenke, K., Williams, J. B. W., & Löwe, B. (2006). A brief measure for assessing generalized anxiety disorder: The GAD-7. Archives of Internal Medicine, 166(10), 1092–1097. https://doi.org/10.1001/archinte.166.10.1092

Stice, E., & Agras, W. S. (1998). Predicting onset and cessation of bulimic behaviors during adolescence: A longitudinal grouping analysis. Behavior Therapy, 29(2), 257–276. https://doi.org/10.1016/S0005-7894(98)80006-3

Stice, E., & Whitenton, K. (2002). Risk factors for body dissatisfaction in adolescent girls: a longitudinal investigation. Developmental Psychology, 38(5), 669–678. https://doi.org/10.1037/0012-1649.38.5.669

Struk, A. A., Carriere, J. S. A., Cheyne, J. A., & Danckert, J. (2017). A Short Boredom Proneness Scale: Development and Psychometric Properties. Assessment, 24(3), 346–359. https://doi.org/10.1177/1073191115609996

Taranis, L., Touyz, S., & Meyer, C. (2011). Disordered eating and exercise: Development and preliminary validation of the compulsive exercise test (CET). European Eating Disorders Review, 19(3), 256–268. https://doi.org/10.1002/erv.1108

Tennant, R., Hiller, L., Fishwick, R., Platt, S., Joseph, S., Weich, S., … Stewart-Brown, S. (2007). The Warwick-Dinburgh mental well-being scale (WEMWBS): Development and UK validation. Health and Quality of Life Outcomes, 5. https://doi.org/10.1186/1477-7525-5-63

Termorshuizen, J. D., Watson, H. J., Thornton, L. M., Borg, S., Flatt, R. E., Macdermod, C. M., … Bulik, C. M. (2020). Early Impact of COVID-19 on Individuals with Eating Disorders: A survey of ∼1000 Individuals in the United States and the Netherlands. MedRxiv, 2020.05.28.20116301. https://doi.org/10.1101/2020.05.28.20116301

Touyz, S., Lacey, H., & Hay, P. (2020). Eating disorders in the time of COVID-19. Journal of Eating Disorders, Vol. 8. https://doi.org/10.1186/s40337-020-00295-3

Vaglio, J., Conard, M., Poston, W. S., O’Keefe, J., Haddock, C. K., House, J., & Spertus, J. A. (2004). Testing the performance of the ENRICHD Social Support Instrument in cardiac patients. Health and Quality of Life Outcomes, 2. https://doi.org/10.1186/1477-7525-2-24

Weiss, D. S., & Marmar, C. R. (1997). The Impact of Event Scale-Revised. In Assessing Psychological Trauma and PTSD: A Practitioner’s Handbook (pp. 399–411).

Weissman, R. S., Bauer, S., & Thomas, J. J. (2020, May 1). Access to evidence-based care for eating disorders during the COVID-19 crisis. International Journal of Eating Disorders, Vol. 53, pp. 369–376. https://doi.org/10.1002/eat.23279

World Health Organization. (2005). Global Physical Activity Questionnaire Analysis Guide GPAQ Analysis Guide Global Physical Activity Questionnaire (GPAQ) Analysis Guide. Retrieved from http://www.who.int/chp/steps/GPAQ/en/index.html

Zigmond, A. S., & Snaith, R. P. (1983). The Hospital Anxiety and Depression Scale. Acta Psychiatrica Scandinavica, 67(6), 361–370. https://doi.org/10.1111/j.1600-0447.1983.tb09716.x

